# Emotional Adaptation During A Crisis: Decline in Anxiety and Depression After the Initial Weeks of COVID-19 in the United States

**DOI:** 10.1101/2020.12.23.20248773

**Authors:** Anastasia Shuster, Madeline O’Brien, Yi Luo, Matthew Heflin, Dongil Chung, Soojung Na, Ofer Perl, Kaustubh Kulkarni, Vincenzo G. Fiore, Xiaosi Gu

**Affiliations:** Department of Psychiatry, Icahn School of Medicine at Mount Sinai, New York, NY; Nash Family Department of Neuroscience, Icahn School of Medicine at Mount Sinai, New York, NY; Fralin Biomedical Research Institute at VTC, Virginia Tech, Roanoke, VA; Department of Biomedical Engineering, UNIST, Ulsan, South Korea

## Abstract

**Objective:** Crises such as the COVID-19 pandemic are known to exacerbate depression and anxiety, though their temporal trajectories remain unclear. The present study aims to investigate fluctuations in depression and anxiety using COVID-19 as a model crisis.

**Methods:** 1,512 adults living in the U.S. enrolled in this online study on April 2^nd^, 2020 and were assessed weekly for 10 weeks (until June 4^th^, 2020; final n=537). Depression and anxiety were measured using the Zung Self-Rating Depression scale and State-Trait Anxiety Inventory (state subscale), respectively, along with demographic and COVID-related questions. Mixed-effects linear regression models were used to examine factors contributing to longitudinal changes in depression and anxiety.

**Results:** Depression and anxiety levels were high in early April, but declined over time (*F*(9,4824)=17.53, *p*<.001 and *F*(9,4824)=23.35, *p*<.001, respectively). In addition to demographic factors such as sex, age, income, and psychiatric diagnoses, we identified some overlapping and some distinct dynamic factors contributing to changes in depression and anxiety: worsening of weekly individual economic impact of COVID-19 increased both depression and anxiety, while increased seven-day change in COVID-19 cases, social media use, and projected pandemic duration were positively associated with anxiety, but not depression.

**Conclusions:** Alongside evidence for overall emotional adaptation, these findings highlight overlapping (economic), yet distinct (change in COVID-19 cases, social media use, and projected COVID-19 duration) factors contributing to fluctuations in depression and anxiety throughout the first wave of COVID-19. These results provide insight into socioeconomic policies and behavioral changes that can increase emotional adaptation in times of crisis.

## Introduction

In early 2020, the coronavirus disease (COVID-19) devastated the globe with catastrophic health and economic consequences. People faced rapidly rising numbers of cases and deaths, overwhelmed healthcare systems, enormous economic strain, and staggering unemployment rates. All the while, individuals were asked to adhere to social distancing guidelines to reduce chances of viral transmission. Thus, amidst the obvious threat to people’s physical health, the pandemic posed a dangerous risk to *mental health*. Indeed, historical precedence for the mental health consequences of pandemics has been well documented in prior research. Increased suicide rates were observed over the course of the 1918 Influenza pandemic, which racked our social, economic, and medical spheres in ways similar to COVID-19 [1]. Research into mental health during the COVID-19 pandemic indicates that the current crisis is no exception. Worsening mental health conditions in adults have already been reported in the United Kingdom [2], United States [3], and Hong Kong [4].

However, humans also often demonstrate incredible emotional adaptability to new situations, even when faced with prolonged hardship [5]. This is considered a form of resilience, which is defined as “the ability to withstand setbacks, adapt positively, and bounce back from adversity” [6]. Recent research has begun to elucidate the individual-level demographic and behavioral determinants of resilience and emotional adaptation. Particularly, increased resilience against developing depression has been linked to higher social support, familial support [7], and education levels [8], while being female [9] or of low socioeconomic status [10] puts one at a higher risk of developing depression. These individual differences are upheld during crises: following a widespread economic downturn, women and low-income individuals were more likely to develop depressive and anxious symptoms [8]. Other studies have shown that behavioral factors contribute to resilience as well: during the COVID-19 pandemic, a study of an Irish sample showed that emotional wellbeing is positively associated with participation in outdoor activities, and negatively associated with excessive intake of COVID-related social media content [11]. Yet, it remains unclear whether individuals will demonstrate such emotional adaptability during a prolonged crisis such as COVID-19; and if so, what factors might contribute to such adaptation.

Here, we examined the effects of the COVID-19 pandemic on depression and anxiety between April 2^nd^ and June 4^th^, 2020, in a community sample in the U.S. 1,512 participants enrolled in an online study and completed questionnaires every week for a 10-week period. Questions spanned a wide range of topics including self-reported depression and anxiety, subjective feelings and beliefs about COVID-19, and demographic information such as age and socioeconomic status (see **Supplementary Materials** for a complete list). Following data collection, two mixed-effects general linear models were conducted to elucidate the variables contributing to fluctuations in depression and anxiety over the 10-week period. These analyses constitute one of the first investigations into the demographic (i.e., unchanging) and dynamic (i.e., unfolding longitudinally) factors associated with mental health during COVID-19.

## Methods

The present study was part of a large web-based longitudinal study examining mental health and decision-making during the first wave of COVID-19 in the U.S.

### Participants

1,512 participants who met eligibility criteria (age between 18-64, current U.S. resident, >90% study participant approval rating) enrolled in the study on a web-based research platform (www.prolific.co) on April 2nd, 2020 (see **Table 1** for summary of characteristics). Exclusion criteria were: (1) voluntary dropout (638 or 42.2% of participants in total, weekly dropout rate between 7.3-14.4%) (2) duplicated entries (28 or 1.85% of participants), and (3) failure in responding accurately to attention-check questions (e.g., “If you are paying attention, please select ‘most of the time’”) at any time point (309 or 20.44% of participants). The final sample included 537 participants who successfully completed all 10 weeks of data collection (252 females (47%), mean age 36.91±13.68, from 48 states) and passed all data quality checks (**Figure 2A**). Excluded (n=975) and included (n=537) participant cohorts did not differ on demographic factors such as sex and income. The final sample was older (t(1497)=4.22, *p*<.001) and had lower depression and anxiety scores at the first time point (t(1493)=-4.56, *p*<.001, t(1495)=-3.53, *p*<.001, respectively) (see **Supplementary Table S1**). However, the trends of overall depression and anxiety scores of the full sample for each timepoint followed the trend of the final clean sample (**Figure 2B**).

**Table 1.**
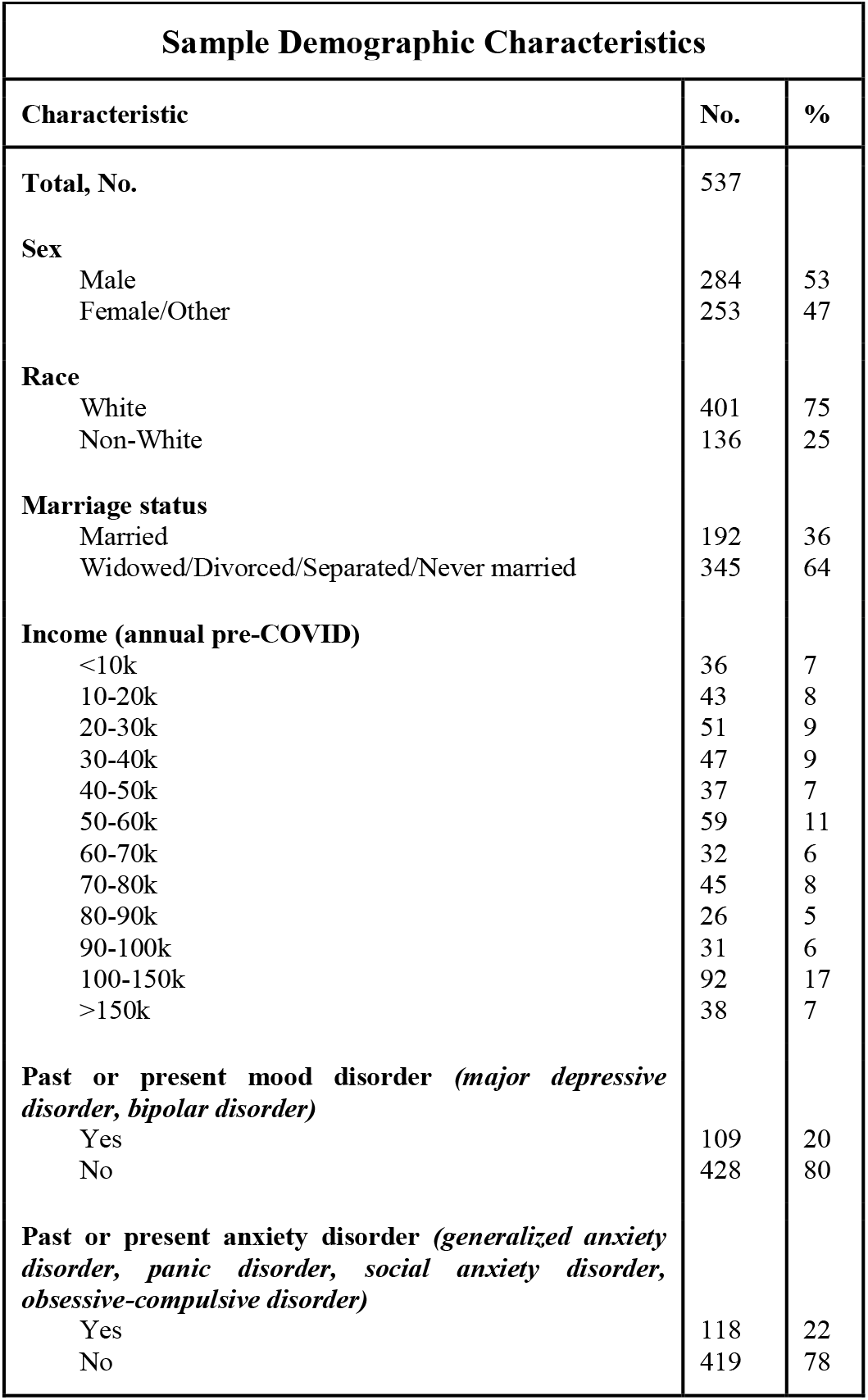
Sample Demographic Characteristics.

| Sample Demographic Characteristics |  |  |
| --- | --- | --- |
| Characteristic | No. | % |
| **Total, No.** | 537 |  |
| **Sex** |  |  |
| Male | 284 | 53 |
| Female/Other | 253 | 47 |
| **Race** |  |  |
| White | 401 | 75 |
| Non-White | 136 | 25 |
| **Marriage status** |  |  |
| Married | 192 | 36 |
| Widowed/Divorced/Separated/Never married | 345 | 64 |
| **Income (annual pre-COVID)** |  |  |
| <10k | 36 | 7 |
| 10-20k | 43 | 8 |
| 20-30k | 51 | 9 |
| 30-40k | 47 | 9 |
| 40-50k | 37 | 7 |
| 50-60k | 59 | 11 |
| 60-70k | 32 | 6 |
| 70-80k | 45 | 8 |
| 80-90k | 26 | 5 |
| 90-100k | 31 | 6 |
| 100-150k | 92 | 17 |
| >150k | 38 | 7 |
| **Past or present mood disorder (*major depressive disorder, bipolar disorder*)** |  |  |
| Yes | 109 | 20 |
| No | 428 | 80 |
| **Past or present anxiety disorder (*generalized anxiety disorder, panic disorder, social anxiety disorder, obsessive-compulsive disorder*)** |  |  |
| Yes | 118 | 22 |
| No | 419 | 78 |

Participants provided informed consent via an online form. The Institutional Review Board of the Icahn School of Medicine at Mount Sinai determined this research to be exempt following review. Participants received base compensation for their time each week ($7.25 for weeks that included behavioral task completion, $3 for weeks that included only survey completion), as well as a scaled bonus according to performance. At week five, participants received a $10 bonus for completing half of the study, and at week 10, participants received a $15 bonus for completing the entire study.

### Procedure

Each week, participants were allotted just over two hours to complete questionnaires assessing mental health as well as perceptions of and behaviors related to COVID-19. Survey data collection was conducted within a 24-hour time window, every 7 days between April 2nd, 2020 and June 4th, 2020 (10 time points in total). Participants additionally provided demographic information at the first time point. Other study elements included decision-making tasks, which are reported elsewhere as they are outside the scope of this study.

### Measures and Scoring

The full survey, as seen by participants at the first time point, can be found in **Supplementary Table S2**. For the analysis, we considered both static demographic factors (e.g., sex) and dynamic factors (fluctuating over time) as variables of interest in relation to mental health.

The initial subset of demographic variables included sex, age, pre-COVID income level (binned into 12 discrete categories), a self-reported history of either a mood or an anxiety disorder diagnosis, and marital status. Marital status was not included in the final analysis due to covariance with other variables of interest (see **Supplementary Information** for details).

Alongside demographic details, answers to the following items were considered in the analysis (**Figure 1B1-3**): (1) economic impact (“Rate the impact that COVID-19 has had on your economic situation”, rated from Very negative impact, −50 to Very positive impact, +50), (2) being informed (“How well are you keeping up with COVID-19 news?” rated from Not at all informed, 0, to Extremely well informed, +100), (3) social media use (“How much do you use social media now/during COVID?”, rated from Not at all, 0, to All the time, +100), (4) and subjective projection of the pandemic’s duration (“How long do you think the global pandemic will last?”, 1-2 months, 3-4 months, 5-6 months, 6-12 months, or More than one year).

**Figure 1.**
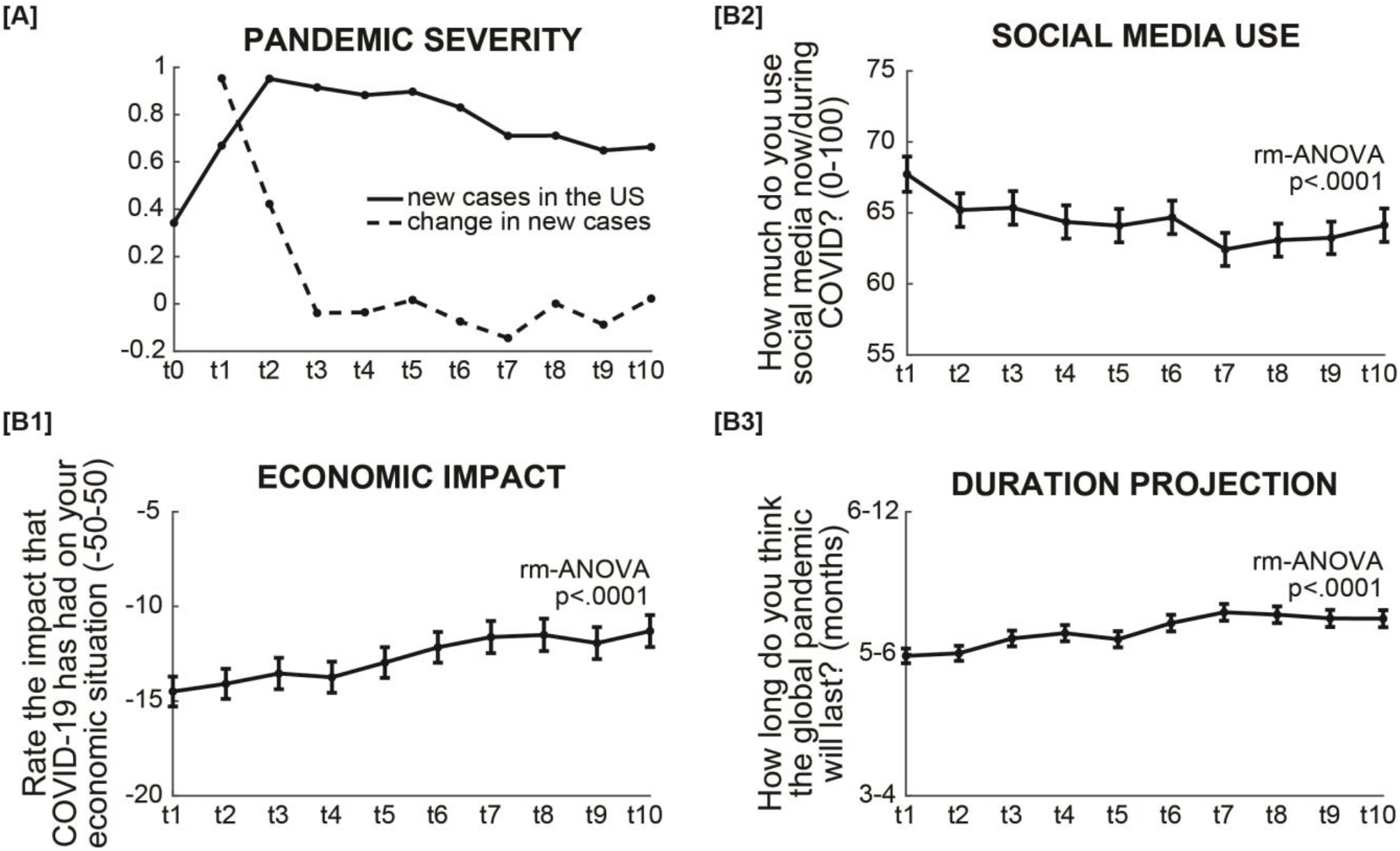
Dynamic variables in the study. **[A]** COVID-19 severity was measured as a 7-day running average of the number of daily new cases (per 10K), and a weekly change in said average between timepoints. **[B]** Self-reported economic impact, social media frequency use, and subjective projection of pandemic’s duration during data collection. Economic impact is overall negative, but improves with time [B1]. Individuals reported using social media less frequently with time [B2]. Individuals’ projected duration for the pandemic increased with time [B3]. Error bars represent standard errors.

The Zung Self-Rating Depression scale [12] and State Anxiety Inventory [13] were used to assess depression and anxiety, respectively.

COVID-19 severity in the U.S. was computed as a 7-day running average of new daily cases [14]. We also calculated a 7-day change in COVID-19 cases by taking the national case count at time point *t* minus national case count at *t-1* (one week earlier), divided by national case count at time point *t-1* (**Figure 1A**).

Physical activity was defined as reporting taking a walk or exercising in the preceding week.

### Statistical analysis

Prior to analysis we preprocessed the variables. We binarized sex into male and female (or other, n=1), race into white and non-white, and marital status into married and not married. We created a mood disorder diagnosis variable by identifying participants with a past or present diagnosis of either major depression or bipolar disorder. We created an anxiety disorder diagnosis variable by identifying participants with a past or present diagnosis of either generalized anxiety disorder, panic disorder, social anxiety disorder, or obsessive-compulsive disorder. Economic impact was scaled to be between -.5 and.5; informedness and social media use were scaled to 0-1.

To identify variables relating to depression and anxiety, we conducted two mixed-effect linear regressions:

Depression (or anxiety) ∼ 1 + age + sex + race + income + diagnosis + age:time + sex:time + race:time + income:time + diagnosis:time + time + COVID19_severity + activity + economic_impact + informedness + social_media + COVID19_future + (1 + age:time + sex:time + race:time + income:time + diagnosis:time + time + COVID19_severity + activity + economic_impact + informedness + social_media + COVID19_future | participant)

All analyses were carried out using MATLAB 2018b. Regressions were conducted using the *fitglme* function.

### Comparison with past studies

To address the lack of pre-pandemic baseline levels of depression and anxiety, we relied on previously reported community means [15,16]. We took the reported mean, standard deviation, and sample size (Depression: M=38.1, SD=9.45, N=172; Anxiety: M=37.28, SD=11.24, N=185), and conducted 2-sample t-tests with the first three time points, for each respective score.

## Results

Both depression and anxiety scores were highest at the beginning of the pandemic, and declined over the 10 weeks (repeated-measures ANOVA, *F*(9,4824)=17.53, *p*<.001, partial *η*^2^=0.03 and *F*(9,4824)=23.35, *p*<.001, partial *η*^2^=0.04, respectively; **Figure 2C&D**). Notably, while average depression scores remained below the standard clinical cutoff for a depression diagnosis [15], the average anxiety score at the first time point (40±13.23) matched a widely-used clinical cutoff of 40, indicating that the average participant in our study was clinically anxious in early April 2020 (t(536)=0.006, *p*=.99, one sample t-test against 40) [17]. Moreover, anxiety at week one was higher than previously reported community averages (37.28±11.24) [16], and decreased to match pre-pandemic averages within three weeks (t1: t(720)=2.5, *p*=.01; t2: t(720)=2.48, *p*=.01; t3: t(720)=1.4, *p*=.12; **Figure 2D**). Depression (week one: 39.3±10.7) followed a similar pattern (community average: 38.10±9.45) [15], albeit not statistically significant (t1: t(707)=1.3; t2: t(707)=0.27; t3: t(707)=0.43; all *p*>.19; see **Figure 2C**).

**Figure 2.**
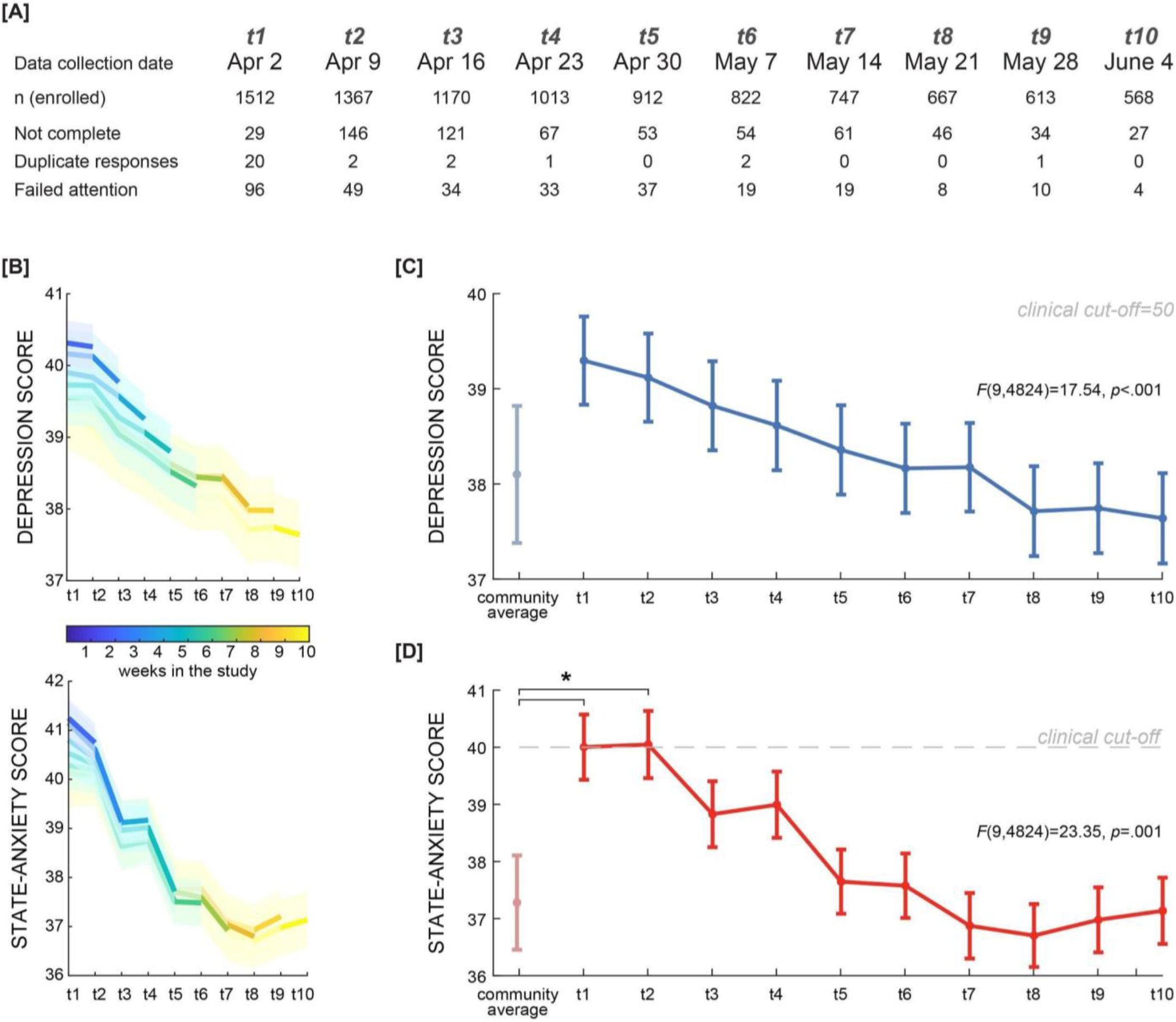
Data collection timeline, with participant exclusion, and depression and anxiety scores. Longitudinal data were collected through weekly surveys for a 10-week period. Final sample included 537 participants. **[A]** Drop-out rates are presented alongside numbers of exclusions based on duplicate responses and failed attention checks. **[B]** Depression and anxiety trends in different subsets of participants. The blue line depicts depression (top panel) and anxiety (bottom panel) scores from participants with valid responses on the first week of data collection (n=1367), who later dropped out or were excluded. The yellow line depicts scores from participants who successfully completed all 10 weeks of data collection (n=537). All other lines depict intermediate subsets of participants. Shaded area represents standard error of the mean. **[C+D]** State-anxiety and depression scores were measured weekly. Error bars indicate standard error of the mean. Community averages were obtained from previously reported depression (light blue) [15] and state-anxiety (pink) [16] scores in healthy adults, with standard error of the mean.

We used two mixed-effects linear models to estimate the influence of demographic and dynamic variables on depression and anxiety separately (which outperformed alternative models consisting of subsets of these variables; see **Supplementary Table S3**). We found similar demographic variables associated with depression and anxiety. Specifically, higher levels of depression and anxiety at each time point were related to being younger (β=-0.2, t(5382)=-7.16, *p*<.001; β=-0.2, t(5382)=-5.48, *p*<.001), female (β=-2.2, t(5382)=-2.74, *p*=.006; β=-1.7, t(5382)=-1.72, *p*=.08), and having lower income (β=-0.6, t(5382)=-4.99, *p*<.001; β=-0.5, t(5382)=-3.73, *p*<.001). As expected, having a diagnosis of a mood or anxiety disorder was also related to a respective increase in depression (β=8.4, t(5382)=8.49, *p*<.001) or anxiety (β=6.7, t(5382)=5.45, *p*<.001) (**Figure 3**).

**Figure 3.**
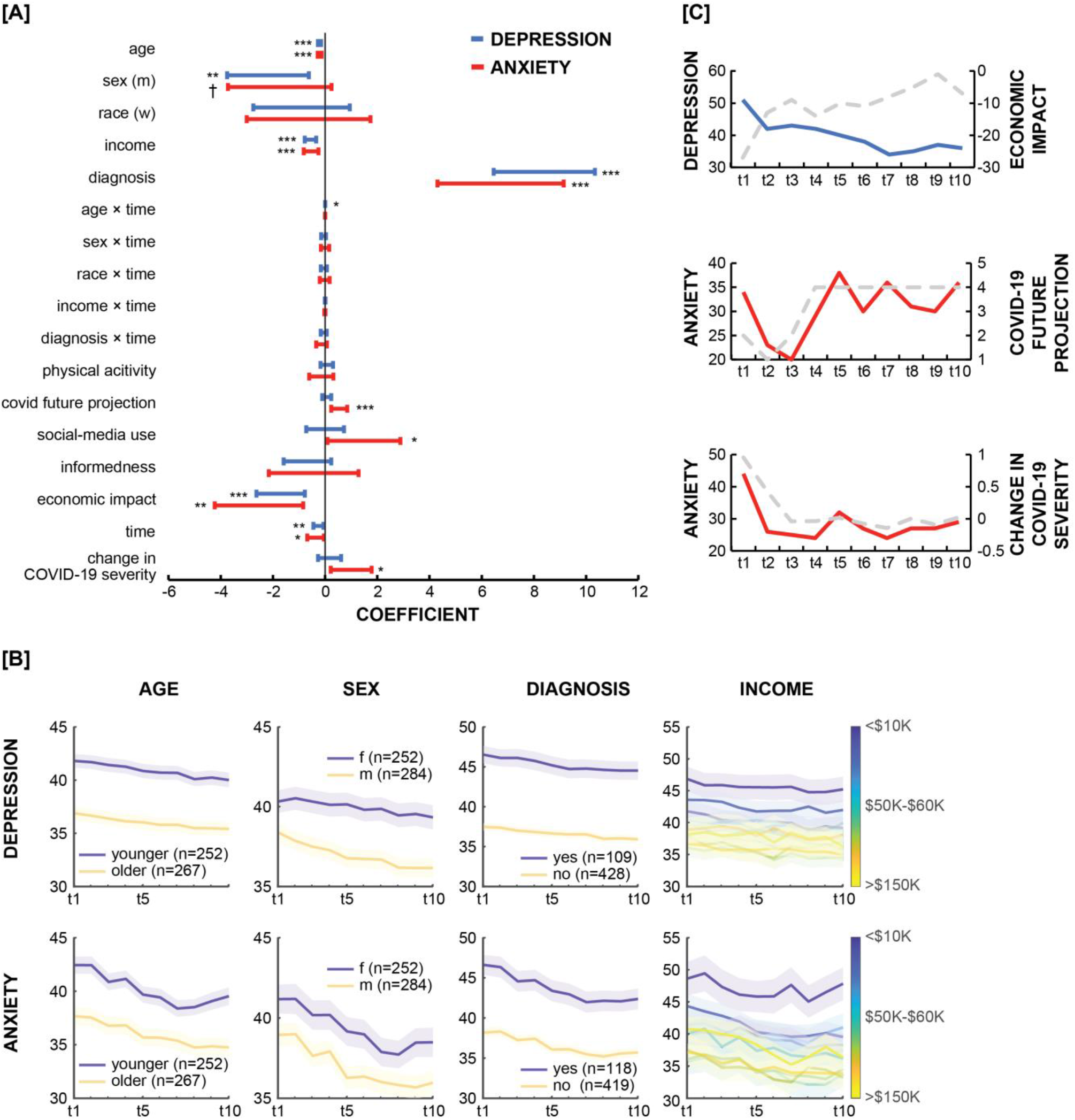
Factors influencing depression and anxiety during the COVID-19 pandemic in the US. **[A]** The coefficients of a mixed-effects linear regression of depression (blue) and anxiety (red). Error bars represent confidence intervals. † *p*<.1, * p<.05, ** p<.01, *** p<.001. **[B]** Illustration of significant demographic variables related to depression and anxiety. **[C]** Example participants depicting significant behavioral and attitude variables related to depression and anxiety. Colored lines represent the example participant’s depression (blue) and anxiety (red). Dotted lines represent the same participant’s behavioral/attitude variable.

For dynamic factors, we found both overlapping and distinct factors predicting changes in depression and anxiety, over and above the effect of time (**Figure 3A**). The economic impact of COVID-19 negatively affected mental health, such that COVID-related worsening of one’s financial situation was related to increases in both depression (β=-1.7, t(5382)=-3.6, *p*<.001) and anxiety (β=-2.5, t(5382)=-2.93, *p*=.003). Interestingly, changes in anxiety, but not depression, were affected by the 7-day change in COVID-19 cases (β=1, t(5382)=2.5, *p*=.01; but not case count itself, **Figure 1A**), social media use (β=1.48, t(5382)=2, *p*=.04), and subjective projections of COVID-19 duration (β=0.54, t(5382)=3.45, *p*<.001). Age interacted with time to predict changes in depression (β=0.004, t(5382)=2.3, *p*=.02) but not anxiety; although older individuals were less depressed overall, time had a less palliative effect on them, as compared with younger individuals. No other demographic factors significantly interacted with time.

## Discussion

Despite the known impact of crises and disasters on mental health, humans are able to adapt to hardship over time [5]. This study provides evidence that in the United States, depression and anxiety initially peaked but then declined over 10 weeks during the first wave of COVID-19. Furthermore, we report that overlapping, yet distinct socioeconomic and psychological factors affected depression and anxiety trajectories as the pandemic lingered. Specifically, the economic impact of COVID-19 negatively affected mental health trajectories, such that COVID-related worsening of one’s financial situation was associated with increases in both depression and anxiety. Changes in anxiety, but not depression, were associated with the 7-day change in national COVID-19 cases, social media use, and subjective projection of COVID-19 duration. Age predicted longitudinal changes in depression but not anxiety, such that the observed decline in depression over time was less strong in older individuals.

Consistent with past work on the link between economic recession and mental health, the economic impact of COVID-19 on individuals was associated with depression and anxiety scores. Following the 2008 Great Recession, a large body of research was conducted to investigate the proximate and long-term emotional ramifications of economic hardship. Financial insecurity and unemployment were found to increase suicide prevalence [18–20], and decrease self-reported happiness and life satisfaction [21]. One investigation into suicide rates in Iceland following the 2008 recession did not find a significant uptick in suicides, a result that the authors attribute in part to “a strong welfare system and investing in social protection” [22]. Income and financial insecurity were also associated with increased depression [20], and psychiatric hospitals reported an increase in outpatient counts among previously healthy individuals, as well as those with anxiety, mood, and adjustment disorders [23]. In the context of COVID-19, complaints of increased self-reported depression and anxiety at the beginning of the pandemic were found to be associated with lower societal appreciation for one’s occupation and the sudden onset of economic hardship [24]. Given the substantial relationship between economic crisis and mental health, especially with regard to income, the field would benefit from future investigations into potential mitigating effects of governmental financial support.

Several variables (7-day change in COVID-19 cases, social media use, and subjective projection of COVID-19 duration) affected anxiety, but not depression. This can potentially be explained by the fundamental differences between depression and anxiety. Historically, scientists have vacillated between consideration of depression and anxiety as the same disorder, different disorders on the same affective spectrum, and entirely different disorders with unique symptomatology [25,26]. Depression and anxiety are very often comorbid in patients with psychiatric disorders [25], and until recently, self-report measures rarely distinguished adequately between their symptoms [27]. But pharmacological separation exists between their treatments (antidepressants and anxiolytics, respectively) [26], and both overlapping and distinct neurobiological profiles have been found to be associated with depression and anxiety in neuroimaging studies [28]. Traditionally, depression has been associated with sadness about the *past*, whereas anxiety has been described as maladaptive anticipation of the *future* [27]. This distinction could, in part, explain why some dynamic factors in the current study measured longitudinally contribute to anxiety and not depression; indeed, the worldwide spread of the coronavirus disease, the prolonged lockdown, and the economic recession have resulted in increased uncertainty concerning one’s future health, as well as interpersonal and financial circumstances. However, such claims are speculative without investigation into the specific differences between depression and anxiety that enabled certain factors to contribute to worsening scores for one disorder but not the other.

In accordance with recent research, we also found that age, sex, and income level were associated with overall depression and anxiety scores such that being younger, female, and having lower income indicated higher scores on average. These results mirror those reported in other studies: a cohort of adults in the United Kingdom who had already completed a mental health study pre-COVID reported higher levels of mental distress during the pandemic, especially young and female participants, in addition to participants with small children [2]. Young adults in the United States were found to have high depression, anxiety, and posttraumatic stress disorder (PTSD) scores during the pandemic, with higher loneliness scores and COVID-related anxiety increasing the likelihood that depression, anxiety, and PTSD scores would reach clinical threshold [29]. In another United States adult sample, depression symptoms were abnormally high at the beginning of the COVID-19 pandemic in comparison to pre-COVID national averages, and individuals with lower income levels reported more depression symptoms than their high-income counterparts [3]. Likewise, self-reported depression and health anxiety symptoms increased particularly rapidly in individuals who experienced financial insecurity in the early days of the pandemic [24]. Thus, the findings reported here provide evidence for individual differences in susceptibility to increased depression and anxiety during COVID-19.

There are two main limitations of the present study. First, no baseline scores for self-reported depression or anxiety could be established due to the unexpected nature of the COVID-19 pandemic and time needed to set up the study. To address this, we compared our scores with previously-reported mean community scores of the same depression and anxiety measures. We found that anxiety and depression both began at higher levels than previously reported community averages - though not significantly so for depression - and decreased to meet these averages by week three of data collection. The second limitation of our study is its correlational nature: with the exception of demographic information and the effects of time and seven-day change of COVID-19 cases, causation in either direction cannot be ascribed to the results of our mixed-effects models. For example, while our findings included an association between increased social media use and exacerbated anxiety scores, this could be explained either by social media content driving anxiety or by the likelihood that an individual with increased anxiety would monitor social media more closely. Likewise, negative economic impact could contribute to depression, but increased depression could also render an individual unable to work. Thus, more research is needed to establish the precise relationships between mental health symptomatology and COVID-19.

Taken together, our findings provide important evidence supporting human resilience as a crisis lingers, as well as revealing the factors contributing to such adaptability. As such, these findings have real-world implications, serving as a vulnerability screening tool in the hands of clinicians and policymakers as they allocate mental health resources during turbulent times. Such a tool would prove most imperative, given the long-lasting effects of COVID-19 on the global economy, as well as other impending crises such as the changing climate.

## Data Availability

Data and analysis code are available upon request.

## Notes

### Competing Interest Statement

The authors have declared no competing interest.

### Funding Statement

No external funding was received for this study.

### Author Declarations

The Institutional Review Board of the Icahn School of Medicine at Mount Sinai determined this research to be exempt following review.

