## Supplementary Materials for "Emotional Adaptation During A Crisis: Decline in Anxiety and Depression After the Initial Weeks of COVID-19 in the United States"

### Supplementary Information

#### Variables exclusion

Prior to fitting the mixed-effects model, we examined the co-dependence between variables. The results are summarized below:

mood-disorder diagnosis & race: χ^2^=8.2, *p*=.004. White participants are more likely than non-white participants to have a mood-disorder diagnosis.

anxiety-disorder diagnosis & race: χ^2^=9.53, *p*=.002. White participants are more likely than non-white participants to have an anxiety-disorder diagnosis.

anxiety-disorder diagnosis & sex: χ^2^=7.41, *p*=.006. females are more likely to have anxiety-disorder diagnosis.

mood-disorder diagnosis & sex: χ^2^=6.96, *p*=.008. females are more likely to have a mood-disorder diagnosis.

sex & income: t(534)=2.1, *p*=.03. Males earn more than females.

age & marriage: t(535)=8.07, *p*<.001. Married individuals are older than unmarried individuals.

race & income: not significant, t(535)=0.8250, *p*=.41.

mood-disorder diagnosis & age: not significant, t(535)=0.04, *p*=.96.

anxiety-disorder diagnosis & age: not significant, t(535)=-0.58, *p*=.56.

mood-disorder diagnosis & marriage: not significant, χ^2^=0.79, *p*=.37.

anxiety-disorder diagnosis & marriage: not significant, χ^2^=3.1715, *p*=.08.

Based on these results, we excluded marital status from the analysis.

### Supplementary Tables

**Table S1. Sample comparisons.** Statistical comparisons between the final sample (n=537) and the participants who either dropped out or were excluded (n=975). Some participants failed to provide responses on some items, and were thus not included in the analysis of those items.

| **Sample comparisons** | | | | | | | | |
| --- | --- | --- | --- | --- | --- | --- | --- | --- |
| **Variable** | **Statistical test** | **Average in included sample (n=537)** | | **Average in excluded sample (n=975)** | | **Statistic (t / z / *X^2^*)** | **DF / rank sum / grand total** | **P value** |
| Depression t1 | two-sample t-test | 39.3 | | 41.9 | | -4.56 | 1493* | 5.3944e-06 |
| Anxiety t1 | two-sample t-test | 40 | | 42.45 | | -3.53 | 1495* | 4.2788e-04 |
| Age | two-sample t-test | 36.9 | | 33.96 | | 4.22 | 1497* | 2.5965e-05 |
| Income | Wilcoxon | 6§ | | 6§ | | 1.18 | 412192† | 0.24 |
|  |  | **n(0)** | **n(1)** | **n(0)** | **n(1)** |  |  |  |
| Sex  0=f/other | chi-square | 253 | 284 | 481 | 481 | 1.149 | 1499‡ | .28 |
| Race  0=non-white | chi-square | 136 | 401 | 232 | 736 | 0.34 | 1505‡ | .56 |
| Marital status  0=not married | chi-square | 345 | 192 | 651 | 317 | 1.4 | 1505‡ | .24 |
| Mood disorder  0=no | chi-square | 428 | 109 | 759 | 209 | 0.35 | 1505‡ | .56 |
| Anxiety disorder  0=no | chi-square | 419 | 118 | 735 | 233 | 0.85 | 1505‡ | .36 |

*: degrees of freedom, †: rank sum, ‡: grand total, §: median

**Table S2. Complete survey.** Questionnaires and survey items as presented to participants during the first data collection time point (April 2, 2020). Bold items were included as independent variables in our analysis.

| **Complete Survey** | | | |
| --- | --- | --- | --- |
| **topic** | **name** | **question** | **options** |
| **demographics** |  | **What is your age?** |  |
|  |  | **What is your sex?** | 1, Male \| 2, Female \| 3, Other |
|  |  | What is your gender identity? | 1, Man \| 2, Woman \| 3, Non-Binary \| 4, Other |
|  |  | **Choose one or more races that you consider yourself to be:** | 1, American Indian or Alaska Native \| 2, Asian \| 3, Black or African American \| 4, Latino or Hispanic \| 5, Multiracial \| 6, Native Hawaiian or Pacific Islander \| 7, White \| 8, Other |
|  |  | If "other", please specify here |  |
|  |  | How many years of school have you received? If you are currently in school, include the current school year. | 1, 1-8 Less than high school \| 2, 9-11 Some high school \| 3, 12 High school degree \| 4, 13-15 Some college \| 5, 16 Bachelor's degree in college (4-year) \| 6, 17-25 Graduate School |
|  |  | **Are you now married, widowed, divorced, separated, or never married?** | 1, Married \| 2, Widowed \| 3, Divorced \| 4, Separated \| 5, Never married |
|  |  | Are you right-handed, left-handed, or ambidextrous? | 1, Right-handed \| 2, Left-handed \| 3, Ambidextrous |
|  | housing situation pre-COVID and change to it | How many people are you currently living with? |  |
|  |  | What is your relationship with these people if you are living with others? | 1, Family \| 2, Friend \| 3, Both \| 4, Roommate \| 5, Myself |
|  |  | Are you living with someone at high risk for contracting COVID-19 (an elderly individual, an immunocompromised individual, etc.) |  |
|  |  | Are you a caregiver to someone in your household? | 1, No \| 2, Child \| 3, Elderly \| 4, Adult(s) with health issues |
|  |  | Are you a caregiver to someone outside of your household? | 1, No \| 2, Child \| 3, Elderly \| 4, Adult(s) with health issues |
|  |  | What type of residence are you living in? | 1, House \| 2, Town home \| 3, Apartment \| 4, Other |
|  |  | If other, please specify. |  |
|  |  | How many bedrooms are in your residence? |  |
|  |  | What is the total size of your residence (in square feet)? |  |
|  |  | Do you have any outdoor space that's part of your residence? | 1, No \| 2, Yes (backyard/front yard) \| 3, Yes (balcony) \| 4, Yes (rooftop) |
|  |  | Have you changed where you live in the past week? |  |
|  |  | How many people are you currently living with? |  |
|  |  | What is your relationship with these people if you are living with others? | 1, Family \| 2, Friend \| 3, Both \| 4, Roommate \| 5, Myself |
|  |  | Are you living with someone at high risk for contracting COVID-19 (an elderly individual, an immunocompromised individual, etc.) |  |
|  |  | Are you a caregiver to someone in your household? | 1, No \| 2, Child \| 3, Elderly \| 4, Adult(s) with health issues |
|  |  | Are you a caregiver to someone outside of your household? | 1, No \| 2, Child \| 3, Elderly \| 4, Adult(s) with health issues |
|  |  | What type of residence are you living in? | 1, House \| 2, Town home \| 3, Apartment \| 4, Other |
|  |  | If other, please specify. |  |
|  |  | How many bedrooms are in your residence? |  |
|  |  | What is the total size of your residence (in square feet)? |  |
|  |  | Do you have any outdoor space that's part of your residence? | 1, No \| 2, Yes (backyard/front yard) \| 3, Yes (balcony) \| 4, Yes (rooftop) |
|  |  | Which political party do you identify with? | 1, Democrat \| 2, Republican \| 3, Independent (lean Democrat) \| 4, Independent (lean Republican) \| 5, Independent (no lean) |
|  |  | Which statement best describes your pre-COVID employment status? | 1, Working (paid employee) \| 2, Working (self-employed) \| 3, Not working (temporary layoff from a job) \| 4, Not working (looking for work) \| 5, Not working (retired) \| 6, Not working (disabled) \| 7, Not working (other) \| 8, Prefer not to answer |
|  |  | If "other", please specify here |  |
|  |  | What is/was your pre-COVID occupation? |  |
|  |  | **Typically (e.g. before COVID), what is your household income level?** | 1, Less than $10,000 \| 2, $10,000 to $19,999 \| 3, $20,000 to $29,999 \| 4, $30,000 to $39,999 \| 5, $40,000 to $49,999 \| 6, $50,000 to $59,999 \| 7, $60,000 to $69,999 \| 8, $70,000 to $79,999 \| 9, $80,000 to $89,999 \| 10, $90,000 to $99,999 \| 11, $100,000 to $149,999 \| 12, $150,000 or more |
|  |  | Think of this ladder as representing where people stand in the United States. At the top of the ladder are the people who are the best off - those who have the most money, the most education, and the most respected jobs. At the bottom are the people who are the worst off - who have the least money, the least education, and the least respected jobs or no job. The higher up you are on this ladder, the closer you are to the people at the very top and the lower you are, the closer you are to the people at the very bottom.  Where would you place yourself on this ladder? | 0, 0 \| 1, 1 \| 2, 2 \| 3, 3 \| 4, 4 \| 5, 5 \| 6, 6 \| 7, 7 \| 8, 8 \| 9, 9 \| 10, 10 |
|  |  | Do you practice any of the following religions? | 1, Baha'i Faith \| 2, Buddhism \| 3, Christianity \| 4, Hinduism \| 5, Islam \| 6, Jainism \| 7, Judaism \| 8, Sikhism \| 9, Taoism \| 10, Rastafarianism \| 11, Not religious \| 12, Other |
|  |  | Which state do you live in? | 1, Alabama \| 2, Alaska \| 52, American Samoa \| 3, Arizona \| 4, Arkansas \| 5, California \| 6, Colorado \| 7, Connecticut \| 8, Delaware \| 9, District of Columbia \| 10, Florida \| 11, Georgia \| 53, Guam \| 12, Hawaii \| 13, Idaho \| 14, Illinois \| 15, Indiana \| 16, Iowa \| 17, Kansas \| 18, Kentucky \| 19, Louisiana \| 20, Maine \| 21, Maryland \| 22, Massachusetts \| 23, Michigan \| 24, Minnesota \| 25, Mississippi \| 26, Missouri \| 27, Montana \| 28, Nebraska \| 29, Nevada \| 30, New Hampshire \| 31, New Jersey \| 32, New Mexico \| 33, New York \| 34, North Carolina \| 35, North Dakota \| 54, North Mariana Islands \| 36, Ohio \| 37, Oklahoma \| 38, Oregon \| 39, Pennsylvania \| 55, Puerto Rico \| 40, Rhode Island \| 41, South Carolina \| 42, South Dakota \| 43, Tennessee \| 44, Texas \| 56, US Virgin Islands \| 45, Utah \| 46, Vermont \| 47, Virginia \| 48, Washington \| 49, West Virginia \| 50, Wisconsin \| 51, Wyoming |
|  |  | Please enter the FIRST THREE digits ONLY of your ZIP code. |  |
| **psychiatric disorders and substance use** | psychiatric disorder diagnosis & medication | Adjustment Disorder | 1, Yes \| 0, No |
|  |  | Attention Deficit Hyperactivity Disorder (ADHD) |  |
|  |  | Autism Spectrum Disorder |  |
|  |  | **Bipolar Disorder** |  |
|  |  | Avoidant Personality Disorder |  |
|  |  | Borderline Personality Disorder |  |
|  |  | **Depression (Major Depressive Disorder - MDD)** |  |
|  |  | Eating Disorder |  |
|  |  | **Generalized Anxiety Disorder (GAD)** |  |
|  |  | **Obsessive Compulsive Disorder (OCD)** |  |
|  |  | Learning Disability |  |
|  |  | **Panic Disorder** |  |
|  |  | Pathological Gambling |  |
|  |  | Post-traumatic Stress Disorder (PTSD) |  |
|  |  | Schizophrenia |  |
|  |  | Schizotypal Personality Disorder |  |
|  |  | **Social Anxiety Disorder** |  |
|  |  | Tourette's Syndrome |  |
|  |  | Substance Use Disorder |  |
|  |  | If you have been diagnosed with another psychiatric disorder, please answer here |  |
|  |  | If you have been diagnosed with a substance use disorder, what is/are your primary substance(s)? | 1, Alcohol \| 2, Nicotine \| 3, Marijuana \| 4, Crack \| 5, Cocaine \| 6, Amphetamine-type stimulants (such as meth) \| 7, Opioid analgesics, including methadone and heroin \| 8, Hallucinogens, including MDMA/ecstasy or mushrooms \| 9, Sedatives and hypnotics, like GHB (excluding benzodiazepine) \| 10, Benzodiazepines, like Xanax \| 11, Inhalants, like spray paint |
|  |  | Do you currently take psychiatric medications? |  |
|  |  | If yes, please list the names of any/all medications here: |  |
|  | substance consumption (pre-COVID) | Alcohol | 0, 0 \| 1, 1 \| 2, 2 \| 3, 3 \| 4, 4 \| 5, 5 \| 6, 6 \| 7, 7 |
|  |  | Tobacco (cigarettes) |  |
|  |  | Tobacco (vaping) |  |
|  |  | Marijuana (traditional methods) |  |
|  |  | Marijuana (vaping) |  |
|  |  | Crack |  |
|  |  | Cocaine |  |
|  |  | Amphetamine-type stimulants (such as meth) |  |
|  |  | Opioid analgesics (including methadone and heroin) |  |
|  |  | Hallucinogens (including MDMA/ecstasy or mushrooms) |  |
|  |  | Sedatives and hypnotics (such as GHB) - excluding benzodiazepines |  |
|  |  | Benzodiazepines (such as Xanax) |  |
|  |  | Inhalants (such as spray paint) |  |
|  |  | Of the days you consumed alcohol per week, how many drinks did you consume per day? |  |
|  |  | Of the days you smoked cigarettes per week, how many cigarettes did you smoke per day? |  |
|  | substance consumption | Alcohol | 0, 0 \| 1, 1 \| 2, 2 \| 3, 3 \| 4, 4 \| 5, 5 \| 6, 6 \| 7, 7 |
|  |  | Tobacco (cigarettes) |  |
|  |  | Tobacco (vaping) |  |
|  |  | Marijuana (traditional methods) |  |
|  |  | Marijuana (vaping) |  |
|  |  | Crack |  |
|  |  | Cocaine |  |
|  |  | Amphetamine-type stimulants (such as meth) |  |
|  |  | Opioid analgesics (including methadone and heroin) |  |
|  |  | Hallucinogens (including MDMA/ecstasy or mushrooms) |  |
|  |  | Sedatives and hypnotics (such as GHB) - excluding benzodiazepines |  |
|  |  | Benzodiazepines (such as Xanax) |  |
|  |  | Inhalants (such as spray paint) |  |
|  |  | Of the days you consumed alcohol in the past week, how many drinks did you consume per day? |  |
|  |  | Of the days you smoked tobacco cigarettes in the past week, how many cigarettes did you smoke per day? |  |
|  | substance craving | In the past week, how much did you crave alcohol? | Not at all \| \| All the time |
|  |  | In the past week, how much did you crave tobacco? |  |
|  |  | In the past week, how much did you crave marijuana? |  |
|  |  | In the past week, how much did you crave crack? |  |
|  |  | In the past week, how much did you crave cocaine? |  |
|  |  | In the past week, how much did you crave amphetamine-type stimulants, such as meth? |  |
|  |  | In the past week, how much did you crave opioid analgesics, including methadone and heroin? |  |
|  |  | In the past week, how much did you crave hallucinogens, including MDMA/ecstasy or mushrooms? |  |
|  |  | In the past week, how much did you crave sedatives and hypnotics, like GHB, excluding Benzodiazepine? |  |
|  |  | In the past week, how much did you crave benzodiazepines, like Xanax? |  |
|  |  | In the past week, how much did you crave inhalants, like spray paint? |  |
| **questionnaires** | state anxiety | I feel calm. | 1, Not at all \| 2, Somewhat \| 3, Moderately so \| 4, Very much so |
|  |  | I feel secure. |  |
|  |  | I am tense. |  |
|  |  | I feel strained. |  |
|  |  | I feel at ease. |  |
|  |  | I feel upset. |  |
|  |  | I am presently worrying over possible misfortunes. |  |
|  |  | I feel satisfied. |  |
|  |  | I feel frightened. |  |
|  |  | I feel comfortable. |  |
|  |  | I feel self confident. |  |
|  |  | I feel nervous. |  |
|  |  | I am jittery. |  |
|  |  | I feel indecisive. |  |
|  |  | I am relaxed. |  |
|  |  | I feel content. |  |
|  |  | I am worried. |  |
|  |  | I feel confused. |  |
|  |  | I feel steady. |  |
|  |  | I feel pleasant. |  |
|  | trait anxiety | I feel pleasant. | 1, Almost never \| 2, Sometimes \| 3, Often \| 4, Almost always |
|  |  | I feel nervous and restless. |  |
|  |  | I feel satisfied with myself. |  |
|  |  | I wish I could be as happy as others seem to be. |  |
|  |  | I feel like a failure. |  |
|  |  | I feel rested. |  |
|  |  | I am calm, cool, and collected. |  |
|  |  | I feel that difficulties are piling up so that I cannot overcome them. |  |
|  |  | I worry too much over something that doesn't really matter. |  |
|  |  | I am happy. |  |
|  |  | I have disturbing thoughts. |  |
|  |  | I lack self-confidence. |  |
|  |  | I feel secure. |  |
|  |  | I make decisions easily. |  |
|  |  | I feel inadequate. |  |
|  |  | I am content. |  |
|  |  | Some unimportant thoughts run through my mind and bother me. |  |
|  |  | I take disappointments so keenly that I can't put them out of my mind. |  |
|  |  | I am a steady person. |  |
|  |  | I get in a state of tension or turmoil as I think over my recent concerns and interests. |  |
|  | UCLS (loneliness) | I lack companionship | 4, "I often feel this way" \| 3, "I sometimes feel this way" \| 2, "I rarely feel this way" \| 1, "I never feel this way" |
|  |  | There is no one I can turn to |  |
|  |  | I am an outgoing person |  |
|  |  | I feel left out |  |
|  |  | I feel isolation from others |  |
|  |  | I can find companionship when I want it |  |
|  |  | I am unhappy being so withdrawn |  |
|  |  | People are around me but not with me |  |
|  | SDS (Zung depression) | I feel down-hearted and blue. | 1, A little of the time \| 2, Some of the time \| 3, Good part of the time \| 4, Most of the time |
|  |  | Morning is when I feel the best. |  |
|  |  | I have crying spells or feel like it. |  |
|  |  | I have trouble sleeping at night. |  |
|  |  | I eat as much as I used to. |  |
|  |  | I still enjoy sex. |  |
|  |  | I notice that I am losing weight. |  |
|  |  | I have trouble with constipation. |  |
|  |  | My heart beats faster than usual. |  |
|  |  | I get tired for no reason. |  |
|  |  | My mind is as clear as it used to be. |  |
|  |  | I find it easy to do the things I used to. |  |
|  |  | I am restless and can't keep still. |  |
|  |  | If you are paying attention, please select "most of the time" |  |
|  |  | I feel hopeful about the future. |  |
|  |  | I am more irritable than usual. |  |
|  |  | I find it easy to make decisions. |  |
|  |  | I feel that I am useful and needed. |  |
|  |  | My life is pretty full. |  |
|  |  | I feel that others would be better off if I were dead. |  |
|  |  | I still enjoy the things I used to do. |  |
|  | OCI (obsessive-compulsive) | I have saved up so many things that they get in the way. | 0, Not at all \| 1, A little \| 2, Moderately \| 3, A lot \| 4, Extremely |
|  |  | I check things more often than necessary. |  |
|  |  | I get upset if objects are not arranged properly. |  |
|  |  | I feel compelled to count while I am doing things. |  |
|  |  | I find it difficult to touch an object when I know it has been touched by strangers or certain people. |  |
|  |  | I find it difficult to control my own thoughts. |  |
|  |  | I collect things I don't need. |  |
|  |  | I repeatedly check doors, windows, drawers, etc. |  |
|  |  | I get upset if others change the way I have arranged things. |  |
|  |  | I feel I have to repeat certain numbers. |  |
|  |  | If you are paying attention, please select 'a little' |  |
|  |  | I sometimes have to wash or clean myself simply because I feel contaminated. |  |
|  |  | I am upset by unpleasant thoughts that come into my mind against my will. |  |
|  |  | I avoid throwing things away because I am afraid I might need them later. |  |
|  |  | I repeatedly check gas and water taps and light switches after turning them off. |  |
|  |  | I need things to be arranged in a particular way. |  |
|  |  | I feel that there are good and bad numbers. |  |
|  |  | I wash my hands more often and longer than necessary. |  |
|  |  | I frequently get nasty thoughts and have difficulty in getting rid of them. |  |
|  | Short dark triad | It's not wise to tell your secrets. | 1, Strongly disagree \| 2, Disagree \| 3, Neither agree nor disagree \| 4, Agree \| 5, Strongly agree |
|  |  | I like to use clever manipulation to get my way. |  |
|  |  | Whatever it takes, you must get the important people on your side. |  |
|  |  | Avoid direct conflict with others because they may be useful in the future. |  |
|  |  | It's wise to keep track of information that you can use against people later. |  |
|  |  | You should wait for the right time to get back at people. |  |
|  |  | There are things you should hide from other people because they don't need to know. |  |
|  |  | Make sure your plans benefit you not others. |  |
|  |  | Most people can be manipulated. |  |
|  |  | People see me as a natural leader. |  |
|  |  | I hate being the center of attention. |  |
|  |  | Many group activities tend to be dull without me. |  |
|  |  | I know that I am special because everyone keeps telling me so. |  |
|  |  | I like to get acquainted with important people. |  |
|  |  | I feel embarrassed if someone compliments me. |  |
|  |  | I have been compared to famous people. |  |
|  |  | I am an average person. |  |
|  |  | I insist on getting the respect I deserve. |  |
|  |  | I like to get revenge on authorities. |  |
|  |  | I avoid dangerous situations. |  |
|  |  | Payback needs to be quick and nasty. |  |
|  |  | People often say I'm out of control. |  |
|  |  | It's true that I can be mean to others. |  |
|  |  | People who mess with me always regret it. |  |
|  |  | I have never gotten into trouble with the law. |  |
|  |  | I like to pick on losers. |  |
|  |  | I'll say anything to get what I want. |  |
|  | subjective happiness | In general, I consider myself: | 1, 1- not a very happy person \| 2, 2 \| 3, 3 \| 4, 4 \| 5, 5 \| 6, 6 \| 7, 7- a very happy person |
|  |  | Compared to most of my peers, I consider myself: | 1, 1- less happy \| 2, 2 \| 3, 3 \| 4, 4 \| 5, 5 \| 6, 6 \| 7, 7- more happy |
|  |  | Some people are generally very happy. They enjoy life regardless of what is going on, getting the most out of everything. To what extent does this characterization describe you? | 1, 1- not at all \| 2, 2 \| 3, 3 \| 4, 4 \| 5, 5 \| 6, 6 \| 7, 7- a great deal |
|  |  | Some people are generally very happy. If you are reading this carefully, please leave this question blank. |  |
|  |  | Some people are generally not very happy. Although they are not depressed, they never seem as happy as they might be. To what extent does this characterization describe you? |  |
|  | perceived stress | In the last week, how often have you felt that you were unable to control the important things in your life? | 0, Never \| 1, Almost never \| 2, Sometimes \| 3, Fairly often \| 4, Very often |
|  |  | In the last week, how often have you felt confident about your ability to handle your personal problems? |  |
|  |  | In the last week, how often have you felt that things were going your way? |  |
|  |  | In the last week, how often have you felt difficulties were piling up so high that you could not overcome them? |  |
|  | ACE (childhood trauma) | Did a parent or other adult in the household often or very often swear at you, insult you, put you down, or humiliate you? Or act in a way that made you afraid that you might be physically hurt? | 1, Yes \| 0, No |
|  |  | Did a parent or other adult in the household often or very often push, grab, slap, or throw something at you? Or ever hit you so hard that you had marks or were injured? |  |
|  |  | Did an adult or person at least 5 years older than you ever touch or fondle you or have you touch their body in a sexual way? Or attempt or actually have oral, anal, or vaginal intercourse with you? |  |
|  |  | Did you often or very often feel that no one in your family loved you or thought you were important or special? Or your family didn't look out for each other, feel close to each other, or support each other? |  |
|  |  | Did you often or very often feel that you didn't have enough to eat, had to wear dirty clothes, and had no one to protect you? Or your parents were too drunk or high to take care of you or take you to the doctor if you needed it? |  |
|  |  | Were your parents ever separated or divorced? |  |
|  |  | Was your mother or stepmother: often or very often pushed, grabbed, slapped, or had something thrown at her? Or sometimes, often, or very often kicked, bitten, hit with a fist, or hit with something hard? Or ever repeatedly hit at least a few minutes or threatened with a gun or knife? |  |
|  |  | Did you live with anyone who was a problem drinker or alcoholic or who used street drugs? |  |
|  |  | Was a household member depressed or mentally ill, or did a household member attempt suicide? |  |
|  |  | Did a household member go to prison? |  |
|  | TALC (tight vs loose community) | There are many social norms that people are supposed to abide by in this country. | 1, Strongly disagree \| 2, Moderately disagree \| 3, Slightly disagree \| 4, Slightly agree \| 5, Moderately agree \| 6, Strongly agree |
|  |  | In this country, there are very clear expectations for how people should act in most situations. |  |
|  |  | People agree upon what behaviors are appropriate versus inappropriate in most situations in this country. |  |
|  |  | People in this country have a great deal of freedom in deciding how they want to behave in most situations. |  |
|  |  | In this country, if someone acts in an inappropriate way, others will strongly disapprove. |  |
|  |  | People in this country almost always comply with social norms. |  |
| **COVID-specific behaviors, attitudes, and beliefs** | attitudes toward COVID-19 guidelines | How important is staying home as much as you can? | Not at all important \| \| Extremely important |
|  |  | How important is keeping a safe distance from other people? |  |
|  |  | How important is washing hands often? |  |
|  |  | How important is covering your cough? |  |
|  |  | How important is self-quarantining when sick? |  |
|  | COVID-19 guidelines behavior | In the past week, how many visitors did you have INSIDE your home? |  |
|  |  | In the past week, how many times per day (on average) did you wash your hands? | 1, 0-5 \| 2, 6-10 \| 3, 11-15 \| 4, 16-20 \| 5, 21+ |
|  |  | In the past week, how often did you sanitize your home, delivery boxes, etc.? | Not at all \| \| All the time |
|  |  | In the past week, how often did you touch your face? |  |
|  |  | In the past week, on average how many times did you go out each day? |  |
|  |  | **To do what?** | 1, Work \| 2, Grocery shopping \| 3, Pharmacy/health-related errand \| 4, Get takeout \| 5, Socialize \| 6, Eat/drink in restaurants/bars \| 7, Exercise (outdoors) \| 8, Going to the gym \| 9, Going for a walk \| 10, Grooming (haircut, manicure/pedicure, etc.) \| 11, Other |
|  |  | If other, please specify. |  |
|  |  | In the past week, how many miles did you travel per day (on average)? |  |
|  |  | In the past week, when you did go out, how often did you come within 6 feet of someone else? | Never \| \| Often |
|  |  | In the past week, did you take any public transportation? |  |
|  | emotions towards COVID-19 situation | How angry are you about the current COVID-19 situation? | Not at all angry \| \| Extremely angry |
|  |  | How disgusted are you by the current COVID-19 situation? | Not at all disgusted \| \| Extremely disgusted |
|  |  | How fearful are you of the current COVID-19 situation? | Not at all fearful \| \| Extremely fearful |
|  |  | How happy are you about the current COVID-19 situation? | Not at all happy \| \| Extremely happy |
|  |  | How sad are you about the current COVID-19 situation? | Not at all sad \| \| Extremely sad |
|  |  | How surprised are you by the current COVID-19 situation? | Not at all surprised \| \| Extremely surprised |
|  | employment during COVID-19 | What is your past week household income before taxes? | 1, Less than $200 \| 2, $200 to $399 \| 3, $400 to $599 \| 4, $600 to $799 \| 5, $800 to $999 \| 6, $1,000 to $1,199 \| 7, $1,200 to $1,399 \| 8, $1,400 to $1,599 \| 9, $1,600 to $1,799 \| 10, $1,800 to $1,999 \| 11, $2,000 to $2,999 \| 12, $3,000 or more |
|  |  | What is your current job? | 1, Transport, retail, or wholesale \| 2, Business and other services, finance, or insurance \| 3, Manufacturing, construction, or agriculture \| 4, Hospitality, catering, or leisure services \| 5, Health or social care \| 6, Public sector or education \| 7, Student \| 8, Unemployed \| 9, Looking for employment \| 10, Other |
|  |  | If other, please specify. |  |
|  |  | If you are working, is your role considered "essential" by the government? |  |
|  |  | If considered essential, what is your role? |  |
|  |  | Rate the impact that COVID-19 has had on your economic situation | Very negative impact \| No impact \| Very positive impact |
|  | COVID symptoms, risk, and exposure | Have you shown any symptoms: dry cough, fever, runny nose, respiratory issues? | 1, Yes \| 0, No |
|  |  | Have you been tested for COVID-19? |  |
|  |  | Have you been diagnosed with COVID-19? |  |
|  |  | Has someone you are living with shown symptoms of COVID-19? |  |
|  |  | Has someone you are living with been diagnosed with COVID-19? |  |
|  |  | Have you been/are you under self-quarantine, meaning you are mandated to have no contact with anyone else? |  |
|  |  | Are you at high risk for developing severe symptoms of COVID-19? |  |
|  |  | If yes, what is/are your pre-existing condition(s) putting you at a higher risk? | 1, Aged 65 or older \| 2, Live in a nursing home or long-term care facility \| 3, Chronic lung disease or moderate to severe asthma \| 4, Serious heart condition \| 5, Immunocompromised \| 6, Severe obesity (BMI > 40) \| 7, Diabetes \| 8, Renal failure \| 9, Liver disease \| 10, Pregnant \| 11, HIV/AIDS \| 12, Prolonged use of corticosteroids |
|  | COVID-19 informedness | **How well are you keeping up with COVID-19 news?** | Not at all informed \| \| Extremely well informed |
|  |  | What sources are you using to get your information? | 1, Fox (TV, social media, internet) \| 2, CNN (TV, social media, internet) \| 3, MSNBC (TV, social media, internet) \| 4, International (BBC, Reuter, etc.) \| 5, Local news (ABC, NBC, CBS, etc.) \| 6, Other |
|  |  | If other, please specify. |  |
|  | social media use | How often did you use social media pre-COVID? | Not at all \| \| All the time |
|  |  | **How much do you use social media now/during COVID?** |  |
|  |  | What type of content do you currently engage with on social media? | 100% non-COVID \| \| 100% COVID |
|  |  | How often do you post on social media vs consume social media content? | 100% post \| \| 100% consume |
|  |  | How often do you engage with/consume content from people you know vs strangers on social media? | 100% people you know \| \| 100% strangers |
|  |  | Which social media outlet do you use most often? | 1, Facebook \| 2, Twitter \| 3, Instagram \| 4, TikTok \| 5, Snapchat \| 6, Reddit \| 7, Other |
|  |  | If other, please specify. |  |
|  | attitudes toward COVID-19 | How is your state government handling the COVID-19 pandemic? | Extremely poorly \| \| Extremely well |
|  |  | How is the federal government handling the COVID-19 pandemic? | Extremely poorly \| \| Extremely well |
|  |  | How disastrous do you think COVID-19 is to our society? | No impact at all \| \| Extremely disastrous |
|  |  | **How long do you think the global pandemic will last?** | 1, 1-2 months \| 2, 3-4 months \| 3, 5-6 months \| 4, 6-12 months \| 5, More than one year |

**Table S3. Model comparison results.** Akaike information criteria (AIC) for the three sub-models included in our model comparison analysis, and the full model reported in the main text: 1) contribution of demographic information to depression and anxiety scores; 2) contribution of demographic information and its interactions with time to depression and anxiety scores; and 3) contribution of demographic information, its interactions with time, and the seven-day change in COVID-19 cases to depression and anxiety scores. The full model consisted of demographic information, its interactions with time, seven-day change in COVID-19 cases, and dynamic measures. The lowest scores (in bold) indicate that the model best fits the data.

| **Model Comparison Results** | | | | |
| --- | --- | --- | --- | --- |
|  | **Demographics** | **Demographics + interactions with time** | **Demographics + interactions with time + seven-day change in COVID-19 cases** | **Full model** |
| Depression | 30007 | 29995 | 29831 | **29809** |
| Anxiety | 36703 | 36733 | 36642 | **36583** |
